# Spatiotemporal Trends in Malnutrition-related Hospitalization and Mortality Among Brazilian Children Under Five

**DOI:** 10.1101/2025.09.24.25336561

**Authors:** Victor Nogueira da Cruz Silveira, Alexandra Mello Schmidt, Mariana Carmona-Baez, Ana Karina Teixeira da Cunha França, Alcione Miranda dos Santos

## Abstract

This study investigates annual hospital admissions and deaths due to malnutrition among Brazilian children under five from 2008 to 2024, analyzing spatial and temporal disparities across microregions and states. Using data from Brazil’s Hospital Information System (SIH), we applied a joint Bayesian spatiotemporal model to examine trends and assess five policy scenarios projected through 2030 in the context of Sustainable Development Goal 2 (SDG2): end all forms of malnutrition by 2030. Results reveal persistent regional inequalities, with the North and Northeast bearing the highest burdens, reflecting deep-rooted structural disparities. Key risk factors included pediatric bed availability (RR 1.13, 95% CrI 1.08–1.18), illiteracy, and low income. The National Hospital Care Policy (PNHOSP) contributed to reduced hospitalizations (RR 0.94, 95% CrI 0.89–0.99), but presented a borderline association with higher odds of death (OR 1.26, 95% CrI 0.98–1.58). Projections suggest that, under current conditions, Brazil is unlikely to meet SDG2 by 2030. Targeted investments in pediatric care infrastructure, combined with broader improvements in the social determinants of health, will be essential to mitigate severe malnutrition outcomes and reduce preventable deaths.

---

In this study, we examine hospital admissions and in-hospital deaths linked to malnutrition among Brazilian children under five years of age. Using national data from 2008 to 2024, we model spatial and temporal patterns across microregions and states and examine associations between these outcomes and policy, socioeconomic, and health system characteristics. We also project how independent and combined changes in infrastructure and policy may affect outcomes through 2030. The findings aim to inform national strategies for mitigating the most severe consequences of malnutrition and to evaluate Brazil’s trajectory toward achieving Sustainable Development Goal 2 (SDG2): ending all forms of malnutrition by 2030.

Early childhood malnutrition arises when a child does not receive adequate nutrients for growth and development, whether from too little food, poor diet quality, or illness that disrupts nutrient absorption.^1, 2^ It is a major contributor to child morbidity and mortality, particularly in low- and middle-income countries, where food insecurity, limited healthcare access, and poor sanitation may be prevalent.^2, 3^ When occurring early in life, nutrient deprivation can disrupt healthy development and have lasting effects well into adulthood and, in some cases, across generations.

Stunting is one of the most visible effects of childhood malnutrition. Defined as low height for age, it reflects prolonged nutritional deficiency and is closely linked to impaired cognitive development, and reduced educational attainment and future economic productivity.^4^ Some studies have also linked childhood stunting to a higher risk of chronic conditions, such as diabetes and hypertension.^5, 6^ At the population level, this translates into reduced economic growth and increased dependency ratios.

Beyond stunting, malnutrition weakens immune defenses, increasing the risk of infection, leading to higher rates of hospitalization for preventable diseases and added strain on health systems.^3, 7^ Global estimates suggest that nearly 45% of all deaths in children under five are attributable to malnutrition, either directly or through increased vulnerability to illness.^8, 9, 10^ Despite this, hospitalization due to malnutrition remains underexamined in global and national analyses. Most hospital-based research focuses on cases of malnutrition that emerge during inpatient hospital stays, rather than those that prompt admission.^11^

In Brazil, such malnutrition-related hospitalizations are categorized as “sensitive to primary care,” meaning they should be preventable through early detection, nutritional support, and timely outpatient care.^12^ Their continued occurrence within the population highlights structural gaps in early detection, health promotion, and nutritional surveillance.

Included in the Sustainable Development Goals, established in 2015 by the United Nations, is the ambitious target to reduce childhood stunting by 40% by 2030.^13, 8^ In Brazil, early national trends suggested progress, stunting fell from 9.9% in 2000 to 6.9% in 2015; but the decline has not held. By 2024, rates had risen to 8.9%.^10^ Still, by 2022, almost one-third of municipalities (31.9%) had met the benchmark of less than 1% stunting. Yet in the North and Northeast, rates remained high at 13.6% and 17.2%, respectively.^14^ As these figures exclude more severe outcomes, including hospitalizations and deaths directly caused by malnutrition, the likely understate the true burden.

## Materials and Methods

### Study population and data sources

This ecological spatiotemporal study included Brazilian children under five years of age who were admitted to public or private hospitals between 2008 and 2024, across the 558 Brazilian microregions. We obtained microdata on hospital admissions for malnutrition (ICD-10 codes E40-E46) and associated in-hospital deaths from the Hospital Information System (*Sistema de Informações Hospitalares - SIH*) using the `microdatasus` package.^15^

The SIH is an open-access information system maintained by the Brazilian Ministry of Health (https://datasus.saude.gov.br/acesso-a-informacao/producao-hospitalar-sih-sus/), containing individual-level data on hospital admissions, surgical procedures, and costs, aggregated to the level of postal address codes for the whole of Brazil. Data are anonymized and include information on the patient’s residence and local sociodemographic context, as reported by the treating hospital.

Brazil, the fifth-largest country by area and seventh most populous (216.4 million inhabitants), is organized into 558 microregions, which are clusters of municipalities with similar demographic, economic, and cultural characteristics. Microregions are distributed across 26 states and one Federal District. These states are further grouped into five macroregions: North, Northeast, Central-West, South, and Southeast.

We identified hospital admissions by filtering records with ICD codes E40-E46 (malnutrition). To avoid duplication from repeated admissions, we used homonym status (yes and no), date of birth, residential zip code, and admission/discharge dates to identify likely readmission. Only the first admission for each child was kept. To reduce bias from high-volume urban centers/capitals, analyses are based on the child’s place of residence rather than location of care. We aggregated final admission counts by microregion and year.

We identified associated in-hospital deaths using a binary variable “death,” which indicated whether the child’s hospitalization ended in death (yes or no). For children with multiple admissions, death was recorded if it occurred during the last hospitalization. Given the low frequency, deaths were aggregated at the state level.

Geospatial shapefiles for microregions and states were obtained using the geobr package,^16^ and covariates were merged with microregion/state codes provided by the Brazilian Institute of Geography and Statistics.

### Covariates

We assembled covariates reflecting sociodemographic characteristics, health policy coverage, service access, and hospital infrastructure. Variables were aggregated by microregion or state, depending on the outcome. While some covariates were used for both outcomes, separate matrices were constructed for admission and deaths. Table 1 provides a summary of the variables used in the analysis.

**Table 1:** Description of the covariates used in the analysis of hospital admissions and deaths related to child malnutrition, Brazil, 2008–2024.

| Covariate | Description |
| --- | --- |
| **Hospital admissions** |  |
| Registration in the national Single Registry (%) | Individuals registered / Population in the microregion |
| Illiteracy (%) | Number of individuals with less than 4 years of study / Population in the microregion |
| Average per capita income in Brazilian reais (R\$) | Mean of the total income of the household / number of residents in the microregion |
| Low birth weight (%) | Number of newborns with low birth weight / total number of births in the microregion |
| Sewage system (%) | Number of households with treated sewage / total number of households in the microregion |
| Treated water (%) | Number of households with treated water / total number of households in the microregion |
| Infant mortality rate (%) | Number of children under 5 years that died / total number of children under 5 years in the microregion |
| Neglected diseases | Number of hospital admissions due to neglected diseases / total number of admissions in the microregion |
| Pediatric beds per 1,000 inhabitants | (Number of pediatric beds / Population of children under 5 years in the microregion) × 1,000 |
| Political leaning of the federal government | 1 if year between 2016 and 2022, 0 otherwise |
| Implementation of the National Hospital Care Policy - 2013 (*Política Nacional de Atenção Hospitalar de 2013 - PNHOSP*) | 1 if year ≥ 2013, 0 otherwise |
| **Deaths** |  |
| ICU requirement | 1 if the admission required ICU, 0 otherwise |
| Length of hospitalization | Total duration of the hospitalization in days |
| Average age | Average age in years of the children who died |
| Pediatric beds per 1,000 inhabitants | (Number of pediatric beds / Population of children under 5 years in the state) × 1,000 |
| Implementation of the National Hospital Care Policy - 2013 (*Política Nacional de Atenção Hospitalar de 2013 - PNHOSP*) | 1 if year ≥ 2013, 0 otherwise |
| Implementation of the Brazil Without Hunger (*Brasil sem Fome*) and return of the *Mais Médicos* program | 1 if year ≥ 2023, 0 otherwise |
| Ongoing pandemic | 1 if year between 2020 and 2023, 0 otherwise |

#### A joint spatiotemporal model

We propose a joint hierarchical spatiotemporal model for the number of admissions and deaths across Brazil, allowing the two outcomes to borrow strength from one another, as they are expected to share similar underlying determinants.

#### Modeling hospital admissions

Let 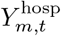 be the number of hospital admissions for microregion *m* (*m* = 1, …, *N*_*m*_) during year *t* (*t* = 1, …, *T*), where *N*_*m*_ = 558 microregions and *T* = 17 years. We assume 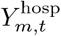, conditional on *λ*_*m,t*_, follows an independent Poisson distribution:

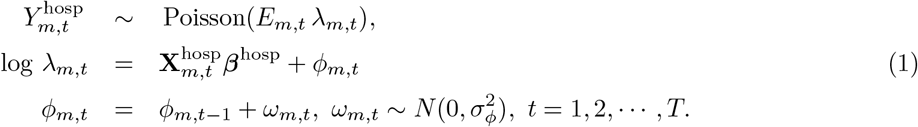

Here *E*_*m,t*_ is an offset term defined as 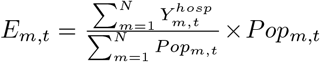, where *Pop*_*m,t*_ denotes the population of children under five years in microregion *m* and year *t*. The term *λ*_*m,t*_ represents the relative risk (RR) of microregion *m* in year *t*. The log-RR is modeled as a unknown linear combination of the covariates and a latent term, *ϕ*_*m,t*_, which captures residual spatial-temporal variation after adjusting for observed covariates. *N* (*µ, σ*^2^) denotes a normal distribution with mean *µ* and variance *σ*^2^.

The latent effect evolves over time via a random walk prior. At the initial time point, *t* = 0, we assume ***ϕ***_0_ = (*ϕ*_1,0_, …, *ϕ*_*N*,0_)′ follows a Leroux prior:^17^

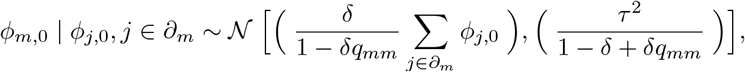

where *∂*_*m*_ denotes the neighboring locations of area *m, q*_*mm*_ represents the number of neighbors of *m*, and *δ ∈* [0, 1] controls the strength of the spatial correlation at *t* = 0. When *δ* = 1, this reduces to a conditional autoregressive (CAR)^18^ prior; when *δ* = 0, *ϕ*_*m*,0_ are independent *a priori*. Substituting *ϕ*_*m,t*_ recursively into equation (1) shows that the latent effect is composed of the initial spatial structure (if there is any) and accumulated temporal noise terms, *ω*_*m,t*_.

#### Modeling the number of deaths among admitted

Because the number of deaths is very small relative to the number of microregions, we aggregate observations to the state level to avoid sparsity. Let 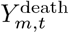 denote the number of children who died after hospitalization in microregion *m* and year *t* and define the number of deaths in state *s* and year *t* as 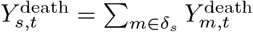, where *s* = 1, 2, …, *N*_*s*_ = 27 and *δ*_*s*_ denotes the set of microregions comprising state *s*. Conditional on the total number of hospital admissions in state *s* and year *t*, 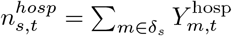, we model the number of deaths as

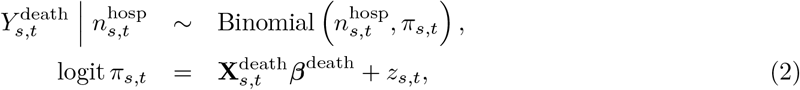

where *z*_*s,t*_ follows a normal prior with mean 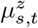 and unknown variance 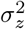.

To encourage information sharing between the admission and death models, we center *z*_*s,t*_ on the average latent effects from the admission model: 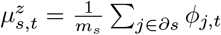, where *∂s* is the set of microregions in state *s*, and *m*_*s*_ is the number of microregions in state *s*.

We adopt a Bayesian approach to estimate the model parameters. Let 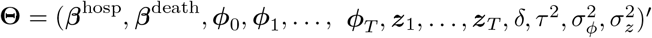 denote the full vector of unknown parameters. We assume prior independence among ***β***^hosp^, ***β***^death^, *ρ, τ* ^2^, 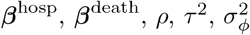, and 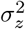. For the regression coefficients, we assign standard normal priors, which provide a plausible range for RRs and odds ratios (ORs).^19^ Scale parameters are assigned half-Cauchy priors with location 0 and scale 1. The spatial dependence parameter *δ*, arising from the Leroux prior, is assigned a uniform prior on (0, 1), allowing the data inform the strength of the spatial correlation.

Given these specifications and Bayes’ theorem, the posterior does not have a closed form. We therefore rely on Markov chain Monte Carlo (MCMC) methods to sample from the posterior. Specifically, we use Hamiltonian Monte Carlo as implemented in the `Stan` probabilistic programming language via the R package `cmdstan`.^20^ We ran two chains of 2,000 iterations each, with a warm-up period of 1,500 iterations. Convergence was assessed through trace plots, effective sample sizes, and the potential scale reduction statistic, 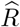.^21^

#### Predicting the number of admissions and deaths six years ahead

To look beyond a simple projection of current trends, we consider five scenarios that reflect both the status quo and possible changes in health infrastructure and policy. These scenarios are built from the available covariates and are used to predict hospital admission and deaths over the next six years. These are described in Table 2.

**Table 2:** Covariate specifications for the five scenarios used to predict hospital admissions and deaths over the six-year period. Covariates not listed are held at their 2024 values.

| Scenario | Description |
| --- | --- |
| 1 | *Status quo* |
| 2 | Pediatric Beds * 1.3 |
| 3 | Pediatric Beds * 0.5 |
| 4 | Pediatric PNHOSP<sup>a</sup>=0 for all $t > 2024$ |
| 5 | PNHOSP=0 for all $t > 2024$ and Pediatric Beds * 2 |

<sup>a</sup> PNHOSP is the National Hospital Care Policy of 2013

Let 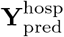 denote the stacked vector of predicted hospital admissions across the *N*_*m*_ microregions, and 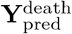 the analogous vector of predicted number of deaths across the *N*_*s*_ states, for years times *T* + 1, …, *T* + 6. Under the Bayesian framework, predictions are obtained through the posterior predictive distribution (PPD), which, under the proposed model, takes the form

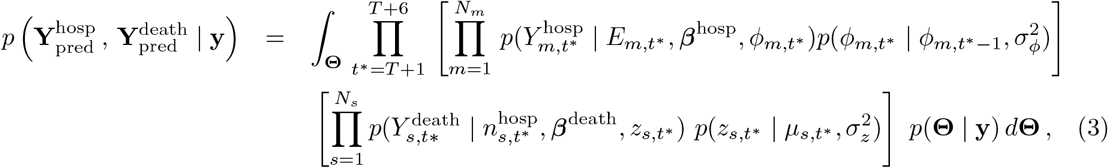

where **y** is the vector containing the observed number of hospitalizations and deaths. A key advantage of the Bayesian framework, as seen in equation 3, is its natural *uncertainty propagation*. Uncertainty in predicted hospital admissions is automatically reflected in the predictions of deaths, as they are modeled jointly. Moreover, uncertainty in the model parameters is fully incorporated into the PPD, yielding more realistic prediction intervals. Because the integral in equation 3 is analytically intractable, we approximate it using posterior samples of **Θ**. Given these samples, draws from the PPD are obtained via a compositional Gibbs sampling procedure, detailed in Appendix S1 of the Supplementary Materials (SM).

#### Ethical aspects

The data in this study come from an open-access information system; therefore, prior requests to government bodies or institutions and approval by the Research Ethics Committee were not required.

## Results

Table 3 summarizes demographic, socioeconomic, and health indicators across Brazil’s macroregions. The North and Northeast have the highest illiteracy rates (15.33% and 24.33%, respectively), the lowest per capita incomes (R$395.05 and R$322.56), and the highest incidence of neglected diseases (over 300 cases per 100,000 inhabitants). In contrast, the Southeast and South show higher per capita incomes (R$693.16 and R$753.89) and greater access to sanitation services (58.84% and 15.03% of treated sewage, respectively). The North and Northeast also account for the highest absolute numbers of hospital admissions and child deaths from malnutrition, while the South and Central-West report the lowest. These patterns vary substantially across states and microregions, motivating our use of spatiotemporal modeling.

**Table 3:**
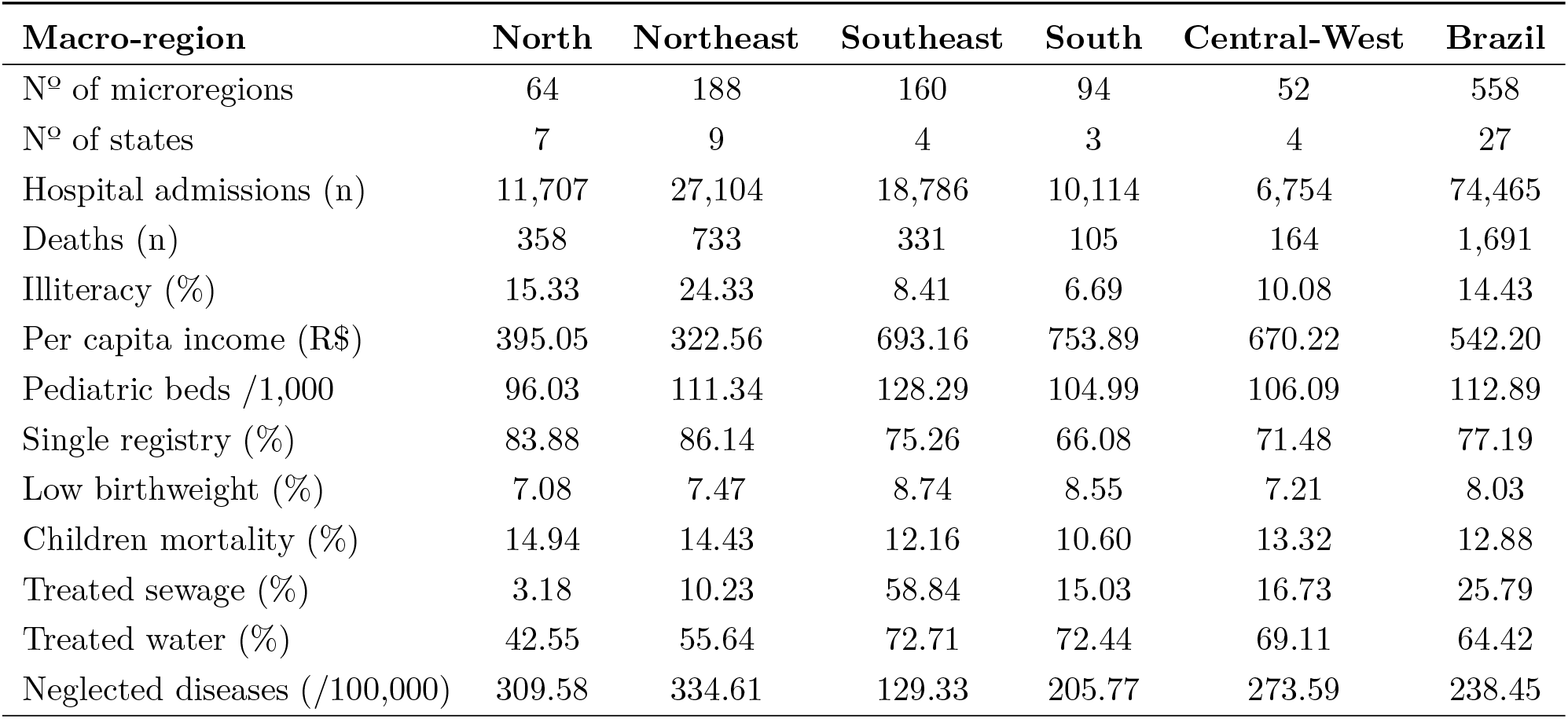
Descriptive statistics of hospital admissions, subsequent deaths, and covariates across Brazil and macro-regions.

The RR of hospital admissions is increased by 13% (RR 1.13; CrI95% 1.08 - 1.18) for each additional pediatric bed per 1,000 inhabitants. A one-unit increase in average income or illiteracy was associated with a 2% increase in RR, although the 95% CrI included 1 in both cases. The implementation of the National Hospital Care Policy (PNHOSP) and the election of a right-wing government were associated with a reduction in RR of 6% (RR 0.94; CrI95% 0.89 - 0.99) and 4% (RR 0.96; CrI95% 0.92 - 0.99), respectively (Figure 1A).

**Figure 1.**
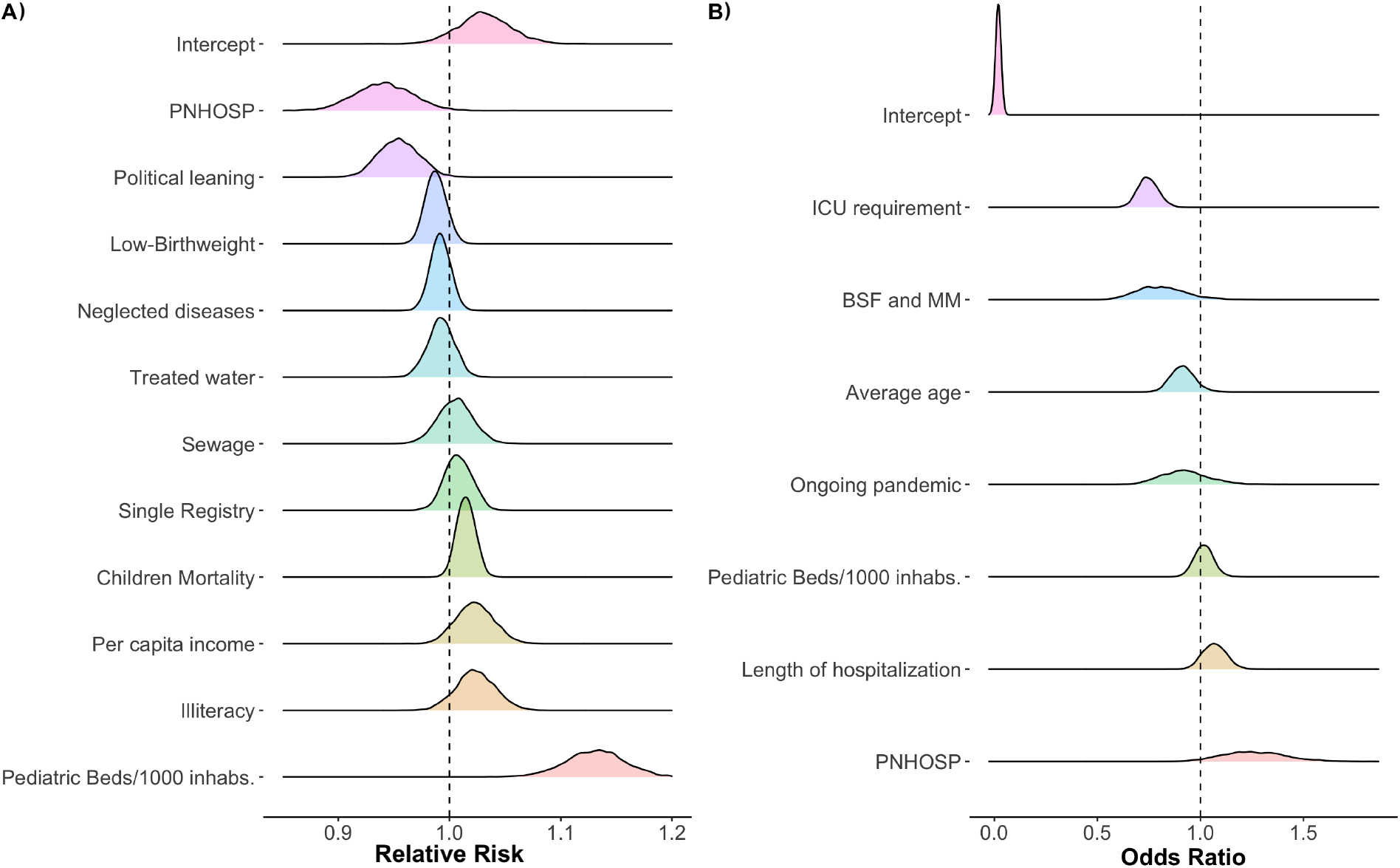
Posterior distribution of the relative risk of hospital admissions (Panel A) and odds ratios of deaths (Panel B) associated with standardized covariates that were included in the respective linear predictors. PNHOSP is the National Hospital Policy Care, ICU is Intensive Care Unit, and BSF and MM are, respectively, *Brasil sem Fome* and *Mais Médicos*

For deaths, admission to intensive care unit (ICU) was associated with a 25% reduction in the odds of death (OR: 0.75; CrI95% 0.66 - 0.85). A one-unit increase in the average age was associated with a 9.1% reduction (OR 0.91; CrI95% 0.81 - 1.04), and the implementation of the *Brasil Sem Fome* (BSF) program and the return of *Mais Médicos*) (MM) in 2023 (OR 0.81; CrI95% 0.61 - 1.07) were also associated with a slight reduction in the odds of deaths. The launch of the PNHOSP in 2013 was borderline associated with a 26% increase in the odds of deaths (OR 1.26; CrI95% 0.98 - 1.58) (Figure 1B).

Figure 2 presents the RR of hospital admissions for selected years (2008, 2016, 2024), illustrating spatial patterns across Brazil. Figure 3 provides a more detailed view of selected microregions over time, including model-based projections. In Brasília, RR values were low or inconclusive until 2020, but increased sharply around 2021 and are projected to remain high, especially under Scenarios 4 and 5 (where PNHOSP = 0). A similar upward trend is seen in Campanha Central and Campanha Meridional, where initially low or moderate risks give way to persistently higher RR in recent years and in projections. In Aglomeração Urbana de São Luís and Altamira, RR values remained consistently high across the observed period and are expected to stay elevated under most scenarios. In contrast, microregions in Ceará and Piauí (Northeast) have maintained low RR over time, a pattern that remains stable across all future projections. Scenario 2, which increases hospital bed capacity, flattens RR trajectories in most areas. The full temporal evolution of RR and the posterior probability of RR exceeding 1 can be seen in the SM (Video S1).

**Figure 2.**
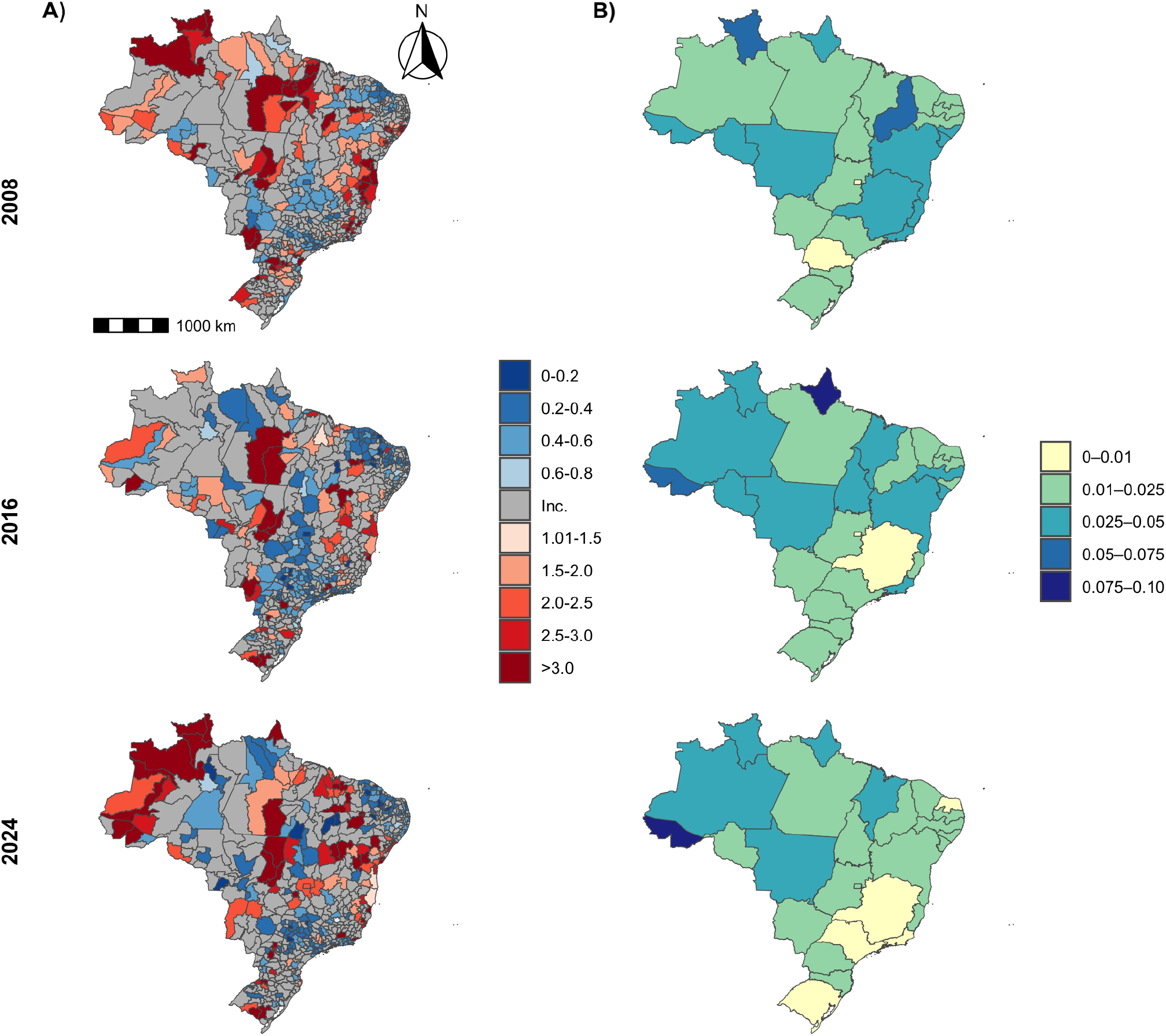
Posterior mean of the relative risk of hospital admissions across microregions (Panel A) and of the probability of deaths (Panel B) across states in 2008, 2016, and 2024 (rows). The legend in the first column is colored according to the 95% posterior credible interval being strictly negative (shades of blue), inconclusive (gray indicated by ‘Inc.’ when 1 is within the 95%CrI), and strictly positive (shades of red).

**Figure 3.**
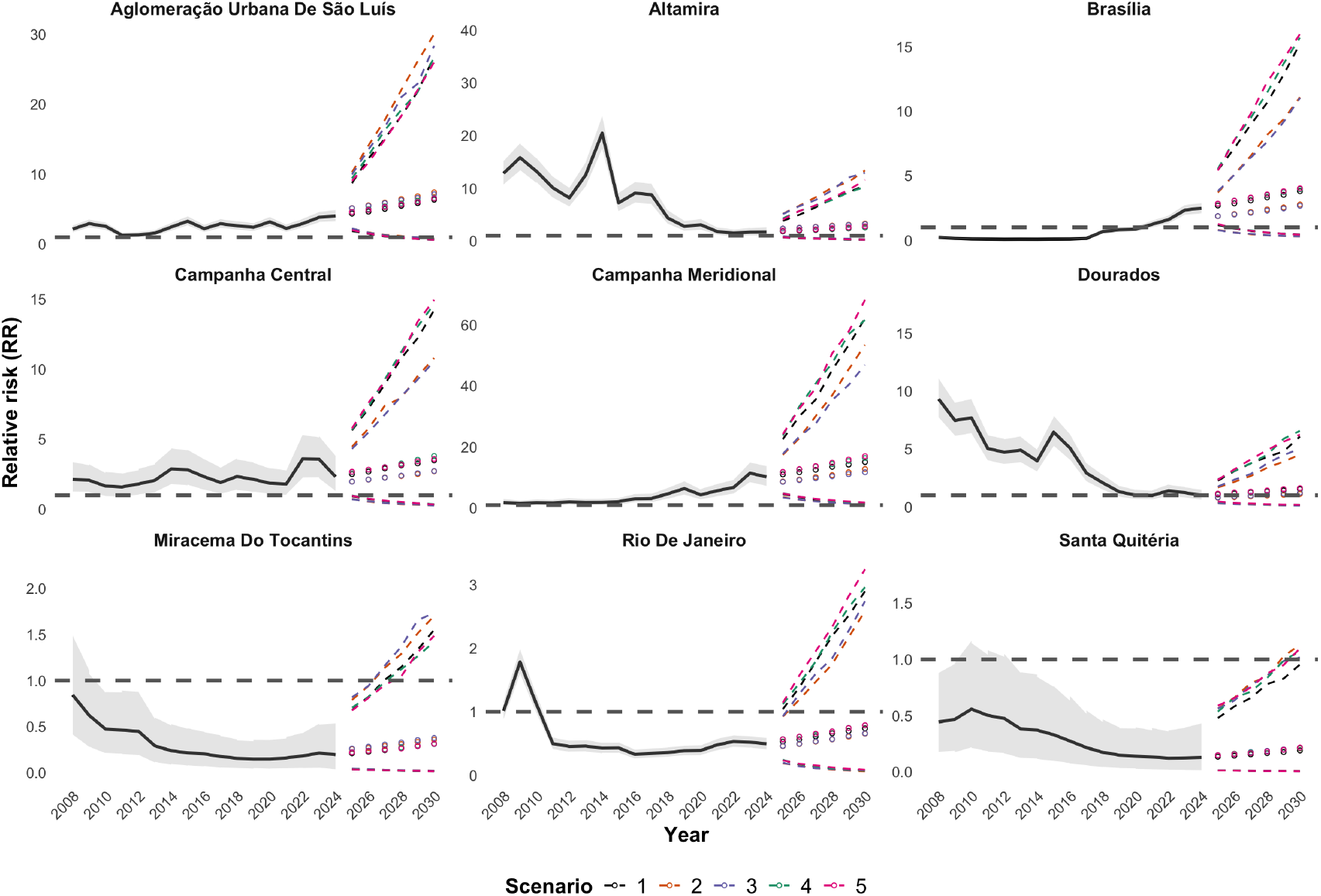
Posterior summaries of estimated (2008-2024) and predicted (2025-2030) relative risks for nine microregions (names on the top of each panel). Solid black lines are the posterior mean of the relative risk. The gray shaded area corresponds to the 95% posterior credible interval of the relative risk. The colored open circles represent the predictive posterior means under each scenario (see legend). Colored dashed lines are the limits of the 95% posterior predictive intervals. The gray horizontal dashed line represents RR = 1.

Figure 4 shows the posterior mean of the probability of death following hospitalization across selected states, with full temporal trends in the SM Video S2. In Acre and Amapá, mortality probabilities increased steadily throughout the observed period, reaching the highest values in the country by the end of the period. In the Federal District (Brasília), early values were among the lowest, but a gradual rise is observed in recent years, with a continued rise projected under all scenarios. Some states, including Roraima, Maranhão, and Piauí, consistently show mortality probabilities above 0.02 with little variation across scenarios. Scenario 4 tends to slightly reduce estimated mortality, while Scenarios 3 and 5 show modest increases. Differences across scenarios are small relative to the width of the credible intervals.

**Figure 4.**
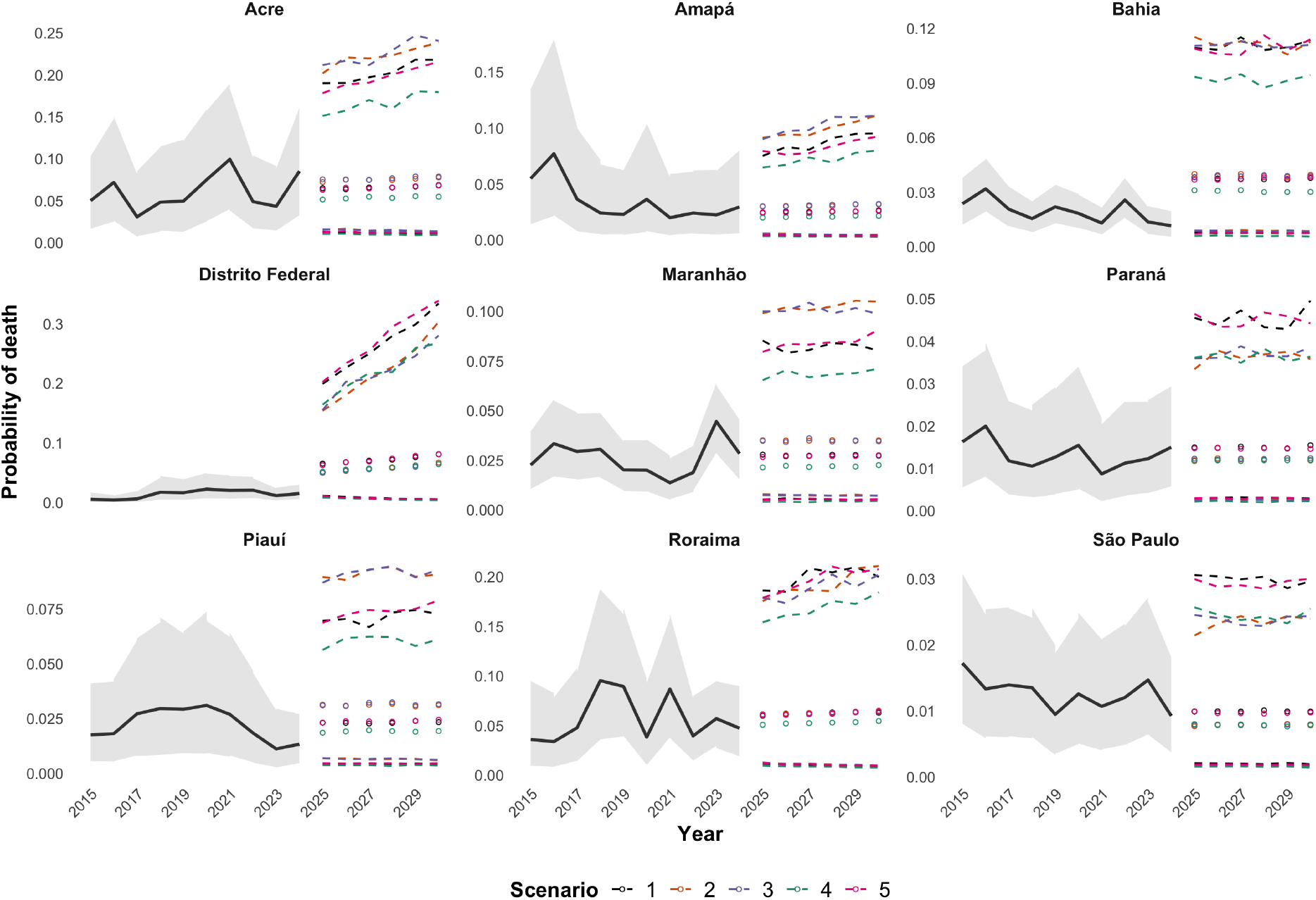
Posterior summaries of estimated (2008-2024) and predicted (2025-2030) probabilities of death for nine states (names on the top of each panel). Solid black lines are the posterior mean of the probability. Gray shaded areas correspond to the 95% credible interval of the estimated probabilities. The colored open circles represent the means of the predictive posterior distribution under each scenario (see legend). Colored dashed lines are the limits of the 95% posterior predictive intervals.

We observed a high probability of a RR *>* 1 in the far west of the North region and in some microregions in Pará (North) (SM Video S1). Microregions in Maranhão (Northeast) consistently exhibited a high risk of hospitalizations across the period. The interior of Bahia (Northeast) and northern Minas Gerais (Southeast) also show a high probability of RR *>* 1 throughout the period. The south of Rio Grande do Sul (South) also shows high risk over the period, although to a lesser extent.

Model diagnostics are provided in the SM, including plots comparing the observed and predicted values for hospital admissions and subsequent deaths due to malnutrition (SM Figures S1 and S2). These results suggest that the model adequately captures the patterns in the data and provides a good fit across the study period.

## Discussion

Hospitalization admissions for malnutrition in children under five are considered avoidable with adequate primary health care (PHC) and are used by the Ministry of Health to monitor PHC performance.^12^ While overall inpatient mortality has declined in Brazil, hospitalizations have remained stable or increased slightly. Given signs of improved care in many areas,^14^ this trend suggests regions with persistent or worsening gaps in early detection and outpatient management.^22^ Our model captures this heterogeneity, identifying clear spatial and temporal patterns in RRs. Our finding that admission rates remain highest in areas with limited PHC coverage but greater pediatric bed availability points to potential substitution of inpatient care where outpatient services are insufficient.

Hospitalization rates varied across regions. For example, starting in 2021, Brasília showed a marked rise in RR, with estimates exceeding 1 and credible intervals excluding 1. This inflection point coincides with the peak of the COVID-19 pandemic in the Federal District, suggesting that the increase may reflect acute stress on the hospital systems and worsening social vulnerabilities.^23^ Bananal, by contrast, maintained low and stable risk throughout, with narrow credible intervals below 1. This trend likely indicates stable demographic and socioeconomic conditions.^24, 25^ Rio de Janeiro showed a small but growing increase with tighter uncertainty, possibly linked to deepening urban inequality and increasing strain on the public health system.^26, 27^

Other regions showed consistent declines in hospitalization rates. In Altamira, the decrease likely reflects population displacement and changing health care demand resulting from the construction of the Belo Monte Hydroelectric Dam,^28^ rather than improved health services. Dourados exhibited a similar trend, but in that case, improvements in the local health system, primary care access, and socioeconomic conditions are a more plausible explanation. However, changes in monitoring or data recording cannot be ruled out.^29^ These contrasts illustrate that similar trends may be driven by different forces, underscoring the importance of local context.

Some of these regional variations may reflect underlying differences in health system factors. Our results show that pediatric bed availability was positively associated with admissions, reflecting either higher burdens of severe malnutrition or delayed care access.^30, 1^ In Brazil, uneven bed distribution, especially in the North and Northeast, may partially explain persistent regional differences in malnutrition mortality.^31, 4^

The relationship between health system factors and outcomes was not uniform. While bed availability was associated with higher admissions, greater access to ICUs was linked to lower mortality: a one-unit increase in the proportion of ICU admissions reduced the odds of death by 25%. This reduction may indicate the survival benefit of timely access to specialized care in severe cases.^32, 33^ In contrast, children with longer hospital stays had higher mortality, suggesting more severe illness at admission or complications during care.^34^

Beyond system-level factors, individual-level social conditions also shaped hospitalization risk. Illiteracy and low income were each modestly associated with higher admissions, consistent with prior evidence linking structural poverty and childhood undernutrition.^5^ Households with limited education and income often experience food insecurity, reduced care access, and difficulty adhering to nutritional guidelines.^35, 36, 37^ Although the estimated effects were small, the high prevalence of socioeconomic vulnerability suggests they may still have meaningful population-level implications.

The broader political context also shaped hospitalization trends. Relative risk of admissions declined during the 2016–2022 right-wing federal administration, but this likely reflects disruptions to surveillance and reporting rather than true improvements.^38, 39^ Cuts to PHC funding and weakened coordination with hospitals may have resulted in reduced outreach and compromised data quality, particularly in poorer regions.^4, 38^

Policy-specific effects were also evident. In 2013, the PNHOSP was introduced to improve integration across care levels and to strengthen referral systems.^22, 40^ Consistent with those goals, it was associated with a reduction in hospitalizations. However, it was also linked to a 26% increase in the odds of death. One possible explanation for these trends is that PNHOSP tightened referral criteria without improving early detection through PHC.^40, 41^ As a result, some children may have reached inpatient care later in the clinical course, increasing the risk of death. Another hypothesis is that although PNHOSP standardized protocols, it did not address persistent gaps in nutritional rehabilitation and follow-up, particularly in remote regions.

In contrast, physician deployment and food security initiatives launched in 2023, (*Brasil Sem Fome* and *Mais Médicos*), were associated with reductions in mortality. These results are consistent with prior evaluations of similar initiatives, which have demonstrated improvements in early detection and reduced progression to severe malnutrition.^42, 43, 44^ Together, expanded PHC and targeted social support appear to have reduced both access barriers and disease progression.

Projections of hospitalizations due to child malnutrition across Brazilian microregions show broadly stable patterns across scenarios, with small differences in mean estimates but important variations in uncertainty. Scenarios involving the removal of the PNHOSP, alone or combined with increased bed availability, tended to produce wider uncertainty intervals, suggesting less predictability in health outcomes when national policy coordination is weakened.^30, 43, 11^

Although some microregions exhibited modest shifts in projected hospitalization risk across scenarios, most differences were small and often overlapped, indicating a high degree of uncertainty around any scenario-specific effect. While the presence of PNHOSP did not substantially change predicted values, it was associated with greater stability and narrower credible intervals, particularly in low-risk areas.^22^

Scenarios involving only structural changes showed inconsistent effects. Some microregions showed slight reductions in hospitalization risk when more pediatric beds were available, while others showed little change. The absence of a consistent effect suggests that infrastructure changes alone are unlikely to shift hospitalization risk unless embedded in broader, integrated policy frameworks.^30^ The wider uncertainty under specific scenarios highlights the fragility of health systems in some microregions and the need for institutional consistency.^43, 5^

Maintaining policies like PNHOSP may help stabilize referral patterns and promote appropriate utilization of care services, especially when coupled with strategic planning.^43, 22^ Without consistent and targeted investment, particularly in structurally disadvantaged regions, even small policy shifts may increase health risks.^5^ The projections reinforce that malnutrition admissions are not solely biological events but are deeply intertwined with broader social and political structures.^1, 30^

Projections of death among admitted children followed similar patterns. Outcomes varied across policy scenarios, rather than converging toward a single trajectory. This variability reflects persistent inequities in infrastructure, access, and institutional capacity, not just demographics or epidemiology.^11^

Again, the removal of PNHOSP emerged as a critical determinant. In several states, scenarios excluding PNHOSP were associated with the lowest projected probabilities of death. While this might appear protective, it more likely reflects decreased detection or admission of severely ill children, underdiagnosis, or delayed care, rather than genuine improvements in health outcomes.^45^ Breakdowns in care coordination, referrals, or surveillance and reporting could manifest as declines in projected mortality, particularly in areas where hospital access is already constrained.^5, 11^

PNHOSP likely acts as a stabilizing mechanism, improving timely referral and standardizing care across care levels.^37, 41^ Its removal appears to amplify blind spots, especially in regions with fragile health infrastructure or weak integration.^40^ Thus, mortality reduction in this context should be interpreted cautiously; they may represent gaps in care delivery rather than true reductions in disease burden.

Scenarios adjusting the number of pediatric beds yielded more heterogeneous results. In some states, increased bed availability was linked to reduced mortality risk, consistent with the benefits of timely inpatient care.^30, 44^ In others, particularly where systems are fragmented, expansion was accompanied by rising projected death probabilities.^43^ The results indicate that adding capacity, without concurrent investment in staffing, training, and infrastructure, can place additional demands on health systems or reveal underlying gaps in service delivery.^43, 5, 4^

Together, these findings raise critical questions about Brazil’s ability to achieve SDG2.^8^ Our projections do not show consistent improvement and underscore the role of institutional continuity and investment in shaping reliable outcomes. Reductions in malnutrition-related hospitalizations and deaths appear to depend not only on preserving policies like PNHOSP but also on expanding system-wide capacity, especially in underserved areas. Variations in future projections are shaped by policy decisions and reflect both opportunities for progress and the risk of setbacks.

This study applies a hierarchical Bayesian model with spatial and temporal structure to jointly analyze hospitalizations and deaths due to malnutrition in Brazilian children under five. It incorporates policy scenarios to forecast progress toward SDG2. Although the approach depends on administrative data that may underreport cases, does not account for all possible confounders, such as food insecurity, and assumes stable future trends, it provides valuable insights for health planning and policy evaluation.

Finally, our analysis points to the need for sustained political commitment to integrated, equitable health policy that seeks to do more than simply scale infrastructure. Attainment of meaningful reductions in hospitalizations and deaths due to malnutrition demands a comprehensive approach to health system governance, with attention to care quality, coordination, and local adaptability. Our projections show that without such a comprehensive approach, Brazil may not meet its global commitment to end malnutrition and prevent avoidable childhood deaths.

## Data Availability

All data produced in the present work are contained in the manuscript

## Acknowledgements

The authors would like to thank the Brazilian Federal Foundation for Support and Evaluation of Graduate Education (CAPES) (finance code 001) for its support in financing the doctoral internship abroad of the author VNCS, which culminated in the development of this work. Schmidt is grateful for financial support from the Natural Sciences and Engineering Research Council (NSERC) of Canada (Discovery Grant RGPIN-2024-04312). We also thank Hannah Doyle for some helpful insights during the preparation of this manuscript.

