## Supplemental Materials for "Spatiotemporal Trends in Malnutrition-related Hospitalization and Mortality Among Brazilian Children Under Five"

*Contents*

Appendix S1: High-level structured algorithm to obtain samples from the posterior predictive distribution for the number of hospital admissions and deaths between 2025 and 2030

Video S1. Spatiotemporal evolution of hospital admission risk due to malnutrition in children under five years across Brazilian microregions from 2008 to 2024. Each frame presents two columns of maps: (A) the posterior probability that the incidence risk exceeds 1, and (B) the estimated incidence risk. Brazil, 2008 – 2024.

Video S2. Spatiotemporal evolution of deaths given admissions due to malnutrition in children under five years across Brazilian states from 2008 to 2024. Brazil, 2008 – 2024.

Figure S1. Counts (2015 to 2024) and forecasts (2025 to 2030) of hospital admissions due to malnutrition in children under five years in Brazilian microregions under different scenarios. Solid black lines are the posterior means of the fitted values obtained after fitting the proposed model. The gray shaded area corresponds to the 95% posterior credible interval. The colored open circles represent the predictive posterior means under each scenario (see legend). The colored dashed lines are the limits of the 95% posterior predictive intervals.

Figure S2. Observed counts (2015 to 2024) and forecasts (2025 to 2030) of deaths given hospital admissions due to malnutrition in children under five years in Brazilian states under different scenarios. The solid black lines represent the posterior mean of the fitted values after fitting the proposed model to the observed data. The gray shaded area corresponds to the 95% posterior credible interval. The colored open circles are the predictive posterior means under each scenario (see legend). The colored dashed lines are the limits of the 95% posterior predictive intervals.

### Appendix S1: High-level structured algorithm to obtain samples from the posterior predictive distribution for the number of hospital admissions and deaths between 2025 and 2030

Assume that a sample of size $L$ is available from the posterior distribution of the parameter vector $\mathbf{\Theta} = (\boldsymbol{\beta}^{\text{hosp}}, \boldsymbol{\beta}^{\text{death}}, \boldsymbol{\phi}_0, \boldsymbol{\phi}_1, \ldots, \boldsymbol{\phi}_T, \boldsymbol{z}_1, \ldots, \boldsymbol{z}_T, \delta, \tau^2, \sigma_\phi^2, \sigma_z^2)'$.

The predictive posterior distribution,

$$p(\mathbf{Y}_{\text{pred}}^{\text{hosp}}, \mathbf{Y}_{\text{pred}}^{\text{death}}) = \int_{\mathbf{\Theta}} \prod_{t^*=T+1}^{T+6} \left[ \prod_{m=1}^{N_m} p(Y_{m,t^*}^{\text{hosp}} \mid E_{m,T}, \boldsymbol{\beta}^{\text{hosp}}, \phi_{m,t^*}) p(\phi_{m,t^*} \mid \phi_{m,t^*-1}, \sigma_\phi^2) \right] \left[ \prod_{s=1}^{N_s} p(Y_{s,t*}^{\text{death}} \mid n_{s,t^*}^{\text{hosp}}, \boldsymbol{\beta}^{\text{death}}, z_{s,t^*}) p(z_{s,t^*} \mid \mu_{s,t^*}, \sigma_z^2) \right] p(\mathbf{\Theta} \mid \mathbf{y})\, d\mathbf{\Theta}\,, \quad (1)$$

can be approximated through Monte Carlo integration,

$$p(\mathbf{Y}_{\text{pred}}^{\text{hosp}}, \mathbf{Y}_{\text{pred}}^{\text{death}}) \approx \frac{1}{L} \sum_{l=1}^{L} \prod_{t^*=T+1}^{T+6} \left[ \prod_{m=1}^{N_m} p(Y_{m,t^*}^{\text{hosp}^{(l)}} \mid E_{m,T}, \boldsymbol{\beta}^{\text{hosp}^{(l)}}, \phi_{m,t^*}^{(l)}) p(\phi_{m,t^*}^{(l)} \mid \phi_{m,t^*-1}^{(l)}, \sigma_\phi^{2^{(l)}}) \right] \left[ \prod_{s=1}^{N_s} p(Y_{s,t*}^{\text{death}^{(l)}} \mid n_{s,t^*}^{\text{hosp}^{(l)}}, \boldsymbol{\beta}^{\text{death}^{(l)}}, z_{s,t^*}^{(l)}) p(z_{s,t^*}^{(l)} \mid \mu_{s,t^*}^{(l)}, \sigma_z^{2^{(l)}}) \right], \quad (2)$$

where

- $p(Y_{m,t^*}^{\text{hosp}^{(l)}} \mid E_{m,T}, \boldsymbol{\beta}^{\text{hosp}^{(l)}})$ is the probability function (pf) of a Poisson distribution with mean, $E_{m,T}\, \lambda_{m,t^*}^{(l)}$, where $\lambda_{m,t^*} = \exp\left(\mathbf{X}_s^{\text{death}(l)} \boldsymbol{\beta}^{\text{hosp}} + \phi_{m,t^*}(l)\right)$ and the offset term, $E_{m,T}$ is fixed at the value for 2024;
- $p(\phi_{m,t^*}^{(l)} \mid \phi_{m,t^*-1}^{(l)}, \sigma_\phi^{2^{(l)}})$ is the pdf of a normal distribution with mean $\phi_{m,t^*-1}^{(l)}$ and variance $\sigma_\phi^{2^{(l)}}$;
- $p(Y_{s,t*}^{\text{death}^{(l)}} \mid n_{s,t^*}^{\text{hosp}^{(l)}}, \boldsymbol{\beta}^{\text{death}^{(l)}}, z_{s,t^*}^{(l)})$ is the pf of the binomial distribution with parameters $n_{s,t^*}^{\text{hosp}^{(l)}} = \sum_{m \in \delta_s} Y_{m,t^*}^{\text{hosp}^{(l)}}$ and probability $\pi_{s,t^*} = \text{expit}\left(\mathbf{X}_s^{\text{death}} \boldsymbol{\beta}^{\text{death}} + z_{s,t^*}\right)$;
- $p(z_{s,t^*}^{(l)} \mid \mu_{s,t^*}^{(l)}, \sigma_z^{2^{(l)}})$ is the pdf of the normal distribution with mean $\mu_{s,t^*}^{(l)}$ and variance $\sigma_z^{2^{(l)}}$.

A sample from the posterior predictive distribution can be obtained by recursively sampling from the distributions above. Algorithm 1 provides a high-level description of how to generate samples from the posterior predictive distribution.

**Algorithm 1** Simulation from the predictive posterior distribution of hospitalizations and deaths, K steps ahead

**for** $l = 1$ **to** $L$ **do**
  **for** $m = 1$ **to** $N_{\text{micro}}$ **do**
    sample $\phi^{(l)}[m, T]$ from $N(\phi^{(l)}[m, T], \sigma_{\phi}^{2^{(l)}})$
    linpred_hosp $\leftarrow \log E_{\text{pred}}[\texttt{hosp\_idx}] + \beta_0^{\text{hosp}} + \mathbf{X}_{\text{hosp\_scen}}[m, T+1]\, \boldsymbol{\beta}^{\text{hosp}^{(l)}} + \phi^{(l)}[m, T+1]$
    sample $Y_{\text{pred}}^{\text{hosp}}[m, T+1]$ from Poisson (exp(linpred_hosp))
    store $\mu^{(l)}[s, T+1] = \sum_{j \in \delta s} \frac{1}{m_s} \phi_{j,T+1}^{(l)}$
    **for** $t = T+2$ **to** $K$ **do**
      sample $\phi_{m,t}^{(l)}$ from $N(\phi_{m,t-1}^{(l)}, \sigma_{\phi}^{2^{(l)}})$
      linpred_hosp $\leftarrow \log E_{\text{pred}}[m, t] + \beta_0^{\text{hosp}} + \mathbf{X}_{\text{hosp\_scen}}[m, t]\, \boldsymbol{\beta}^{\text{hosp}^{(l)}} + \phi_{m,t}^{(l)}$
      sample $Y_{\text{pred}}^{\text{hosp}^{(l)}}[m, t]$ from Poisson (exp(linpred_hosp))
    **end for**
  **end for**
  **for** $s = 1$ **to** $N_s$ **do**
    store $n[s, t]^{(l)} = \sum_{m \in \delta s} Y_{\text{pred}}^{\text{hosp}^{(l)}}[m, t]$
    store $\mu^{(l)}[s, t] = \sum_{j \in \delta s} \frac{1}{m_s} \phi_{j,t}^{(l)}$
    sample $z^{(l)}[s, T+1]$ from $N(\mu^{(l)}[m, T], \sigma_z^{2^{(l)}})$
    prob_death $\leftarrow$ expit $\left(\beta_0^{\text{death}} + \mathbf{X}_{\text{death\_scen}}[m, T+1]\, \boldsymbol{\beta}^{\text{death}^{(l)}} + z^{(l)}[m, T+1]\right)$
    sample $Y_{\text{pred}}^{\text{death}^{(l)}}[m, T+1]$ from Binomial $\left(n^{\text{hosp}^{(l)}}[s, T+1]\ ,\ \text{prob\_death}\right)$
    **for** $t = T+2$ **to** $TK$ **do**
      sample $z^{(l)}[s, t]$ from $N(\mu^{(l)}[m, t], \sigma_z^{2^{(l)}})$
      prob_death $\leftarrow$ expit $\left(\beta_0^{\text{death}} + \mathbf{X}_{\text{death\_scen}}[m, t]\, \boldsymbol{\beta}^{\text{death}^{(l)}} + z^{(l)}[m, t]\right)$
      sample $Y_{\text{pred}}^{\text{death}^{(l)}}[m, t]$ from Binomial $\left(n^{\text{hosp}^{(l)}}[s, t]\ ,\ \text{prob\_death}\right)$
    **end for**
  **end for**
**end for**

Hospital admission video

Death video

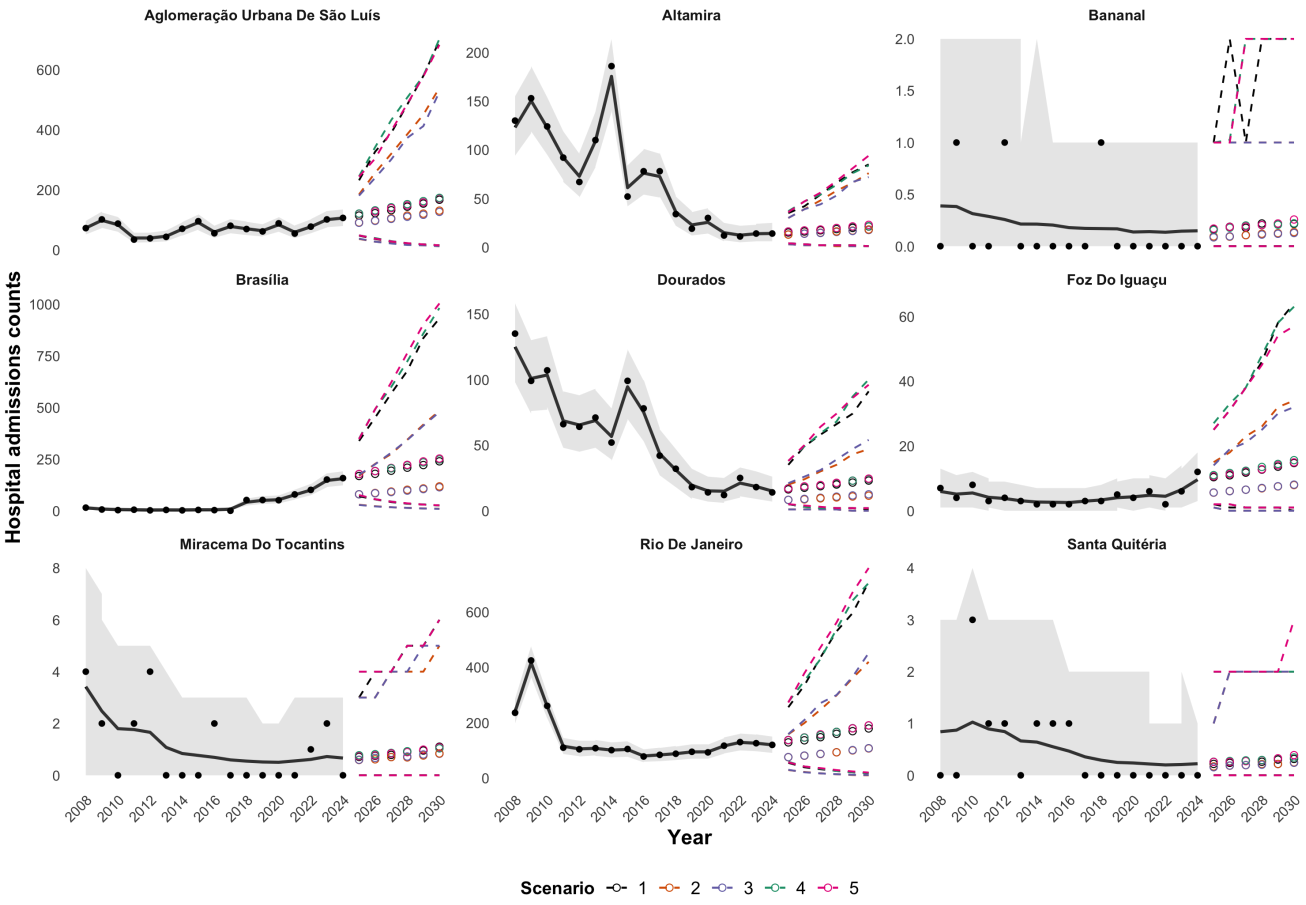


Figure 1: Counts (2015 to 2024) and forecasts (2025 to 2030) of hospital admissions due to malnutrition in children under five years in Brazilian microregions under different scenarios. Solid black lines are the posterior mean of the counts obtained from the observed data. Solid black circles are observed values. Gray shaded area corresponds to the pointwise 95% posterior credible interval of the fitted values. The colored open circles represent the predictive posterior means under each scenario (see legend). The colored dashed lines are the limits of the 95% posterior predictive intervals.

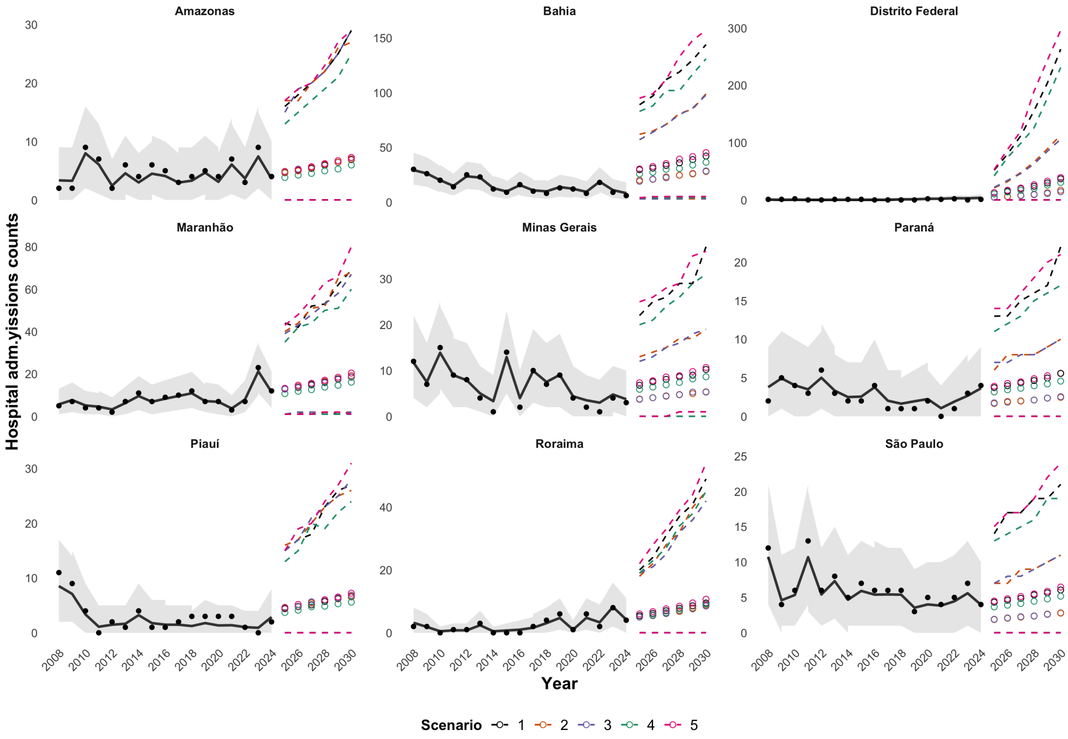


Figure 2: Observed counts (solid black circles from 2008 to 2024) and forecasts (open circles from 2025 to 2030) of deaths given hospital admissions due to malnutrition in children under five years in Brazilian states under different scenarios. Solid black lines are the posterior means of the fitted values obtained after fitting the proposed model. Solid black circles are observed values. Gray shaded area corresponds to the pointwise 95% posterior credible interval of the fitted values. The colored open circles represent the predictive posterior means under each scenario (see legend). The colored dashed lines are the pointwise limits of the 95% posterior predictive intervals.
